# Evaluation of Multifaceted Patient-Peer Delivered Intervention for Type-2 Diabetes Control and Remission in Rural Locations in India: Open-Label Cluster Randomised Pilot Study

**DOI:** 10.64898/2026.03.08.26347876

**Authors:** Krishna Kumar Sharma, Shailendra Kumar B Hegde, Radha Valaulikar, Sowmya Garigipati, Anne-Marie Ernst-Stegeman, Emma Coles, Hanno Pijl, Najeeb Hazarika, Devika Gali, Manisha Choudhury, Anupa Vig, Chiranjib Baruah, Ramnath Ballala, Alexander C Boers, Luke Bredius, Gracia Habib, Eleftheria Vergou, Hamza Yousuf, Nynke van der Zijl, Sailesh Lodha, Leonard Hofstra, Jan van den Berg, Rajeev Gupta

## Abstract

**Objective:** To evaluate the effectiveness of a multifaceted lifestyle intervention delivered by patient peers and supported by healthcare workers and technology to achieve control and remission of type 2 diabetes.

**Methods:** Open-label, cluster-randomised controlled trial conducted in rural Assam, India. Type 2 diabetes patients identified through a screening program in 25 villages (clusters) were randomly assigned to intervention or standard care. At baseline, all participants underwent assessment of diet, physical activity, quality of life, medicine intake, physical measurements and biochemical evaluation. The intervention was a contextually designed package, delivered during fortnightly group counselling by patient-peers and healthcare workers, focusing on carbohydrate-restricted healthy diet, physical activity, diabetes management, and medication de-escalation. Nutrition data were transferred to the study management for suggestions and modifications on smartphones. Intervention was implemented for 3 months, when anthropometric and biochemical parameters were reassessed. Primary outcomes were diabetes control (HbA1c, fasting glucose) and remission (HbA1c <6.5% without medications). Modified intention-to-treat analysis has been performed.

**Results:** 353 patients in rural locations (intervention=193, standard care=160) were enrolled. Baseline sociodemographic, lifestyle, clinical and biochemical parameters were not different in intervention and standard-care groups. At 3 months, in the intervention vs standard-care group there was significantly lower median (interquartile range) HbA1c 7.9% (7.0-8.7) vs 8.6% (7.6-9.8), p<0.001; and fasting glucose 188.0 mg/dl (146.2-253.5) vs 210.0 mg/dl (166.0-282.0), p=0.001. Diabetes remission was in 9 participants (5.0%) in intervention vs 4 (2.7%) in standard care (p=0.249).

**Conclusions:** Patient-peers delivered and healthcare worker- and technology-supported diet and lifestyle intervention for type 2 diabetes led to significant improvement in diabetes control in rural patients in India. Diabetes remission was observed in a low proportion.

**Trial registration:** Registered with Clinical Trials Registry of India at www.ctri.nic.in; registration number CTRI/2022/03/041302 dated 22 March 2022.

**STRENGTHS AND LIMITATIONS OF THE STUDY:**

- Multifaceted diet and lifestyle intervention for type-2 diabetes control and remission, delivered by patient-peers with health worker and technology support, is feasible in rural populations in India.
- This cluster-randomised trial shows that the intervention led to significantly better diabetes control. Diabetes remission occurred in a small proportion.
- Larger and longer prospective studies are required to confirm the effectiveness of such lifestyle strategies for diabetes control and remission in India, lower-middle, and low-income countries.

## INTRODUCTION

The Global Burden of Disease study has predicted that by the year 2050, there will be more than 1.3 billion patients with type 2 diabetes worldwide, more than double the current 530 million.^1^ Most of this burden is projected in lower-middle and middle-income countries of Asia and Africa.^1^ A pooled survey from 55 low and middle-income countries reported that only one in ten patients receives comprehensive diabetes care.^2^ A national survey conducted by the Indian Council of Medical Research reported a prevalence of 101 million type 2 diabetes patients in India, in addition to 136 million with prediabetes.^3^ It has been reported that more than 80% of type 2 diabetes patients in India are either unaware of the disease or have uncontrolled diabetes and present to the healthcare system with a more advanced condition.^4^ Type 2 diabetes related mortality is significantly greater in India and many lower-income countries compared to higher-income countries,^5^ and the economic burden of diabetes in India and other lower-income countries is substantial.^6^

In India, policy initiatives have prioritised strengthening the health system’s capacity to manage various non-communicable diseases, including type 2 diabetes.^7,8^ Clinical trials conducted mostly in urban areas of the country have reported that comprehensive non-physician health worker-delivered interventions can prevent type 2 diabetes among individuals with impaired glucose tolerance.^9,10,11,12,13^ Studies have reported that the benefits of such interventions include reducing cardiovascular risk in type 2 diabetes.^14,15^ Telehealth initiatives, delivered either directly or remotely, have reported benefits in type 2 diabetes prevention and control,^16,17,18,19,^ including reversal of the metabolic syndrome.^20^ The DiRECT trial, a cluster randomized trial in the UK, reported that dietary counselling using health-worker-led group-based intervention led to significant weight loss, effectively reduced Hb1Ac, and induced type 2 diabetes remission in a third.^21^ The remission persisted in one-third of the participants at 24 months.^22^ However, the long-term benefits of such a strategy have been variable. The UK-National Health Service Type 2 Diabetes Path-to-Remission program reported remission in 12-35% at 12-month follow-up.^23^ Studies in other parts of the world have provided discordant results.^24,25,26,27,28,29^ Building on this evidence, we aimed to test the hypothesis that a lifestyle intervention, tailored to the local culture and healthcare circumstances, and delivered by a patient-peer with healthcare worker and technology support shall improve glycemic control (HbA1c) and induce diabetes remission in rural patients with type 2 diabetes in India.

## METHODS

This is a cluster-randomised controlled parallel-group trial in villages in Assam (India) for patients with type 2 diabetes. The intervention and standard care were randomized at the village (cluster) level. Standard care, or standard care plus lifestyle intervention, was delivered by trained patient-peers supported by healthcare workers. The institutional ethics committees at Piramal Swasthya Management and Research Institute, Hyderabad (Letter No PSMRI/2020/06, dated 8 February 2020) reviewed and approved the study protocol. This study is registered at the WHO-listed Clinical Trials Registry of India (Indian Council of Medical Research) at website: www.ctri.nic.in; registration number CTRI/2022/03/041302, dated 22 March 2022. The urban location of Jaipur (Rajasthan, India) was also identified as a site for the study, ethical clearances were obtained, and the study was initiated. However, due to the COVID-19 epidemic, recruitment was slow and low, and therefore, data from the urban patients have not been processed.

### Participants

Type 2 diabetes patients in multiple villages (rural clusters) in Assam were identified. The inclusion criteria were the presence of clinician-diagnosed type 2 diabetes, on medications, with no major life-threatening co-morbidities, capable of providing informed consent, and being able to measure blood glucose using a glucometer. The exclusion criteria were age less than 18 or more than 65 years, using insulin for diabetes control, recent acute coronary syndrome (less than 3 months), any heart failure, presence of cancer, eating disorder, pregnancy, elevated urinary albumin-to-creatinine ratio, or active chronic disease compromising participation. Participants were required to provide informed consent for participation after reading the information sheet in the local language. Each participant provided informed consent-audio-visual or written consent. The village was considered a cluster, and each cluster was allocated to either the intervention or standard care arm through a simple randomization protocol.

#### Intervention

The intervention targeted various facets of individual lifestyles, including dietary patterns, physical activity, stress management, sleep quality, and gradual reduction of type 2 diabetes medication.^30^ The standard care arm continued their usual treatment, consistent with the national guidelines and practices.^31^ Health services in the rural communities were provided through a public-private partnership project between Piramal Swasthya Management and Research Institute and the Government of Assam, India. During the study period, only the on-site health workers were aware of randomization of the study participants. Follow-up care was delivered by an existing network of healthcare services equipped with a telemedicine facility. The Dutch “ReverseDiabetes2Now” multicomponent lifestyle program, developed by the foundation Voeding Leeft, was adopted and tailored to the local language and culture.^30,32,33^ It was converted to the local context and the Assamese language. The protocol and cuisines were developed in a collaborative process led by Joep Lange Institute,^30^ and carried out by a team consisting of experts from Piramal Swasthya Management and Research Institute, India,^34^ with support from Voeding Leeft.^30^ The lifestyle program and training materials, including a recipe book, were developed in Assamese, explaining the pathophysiology of type 2 diabetes and the effects of nutrition, sleep, relaxation and physical activity on the disease.

The 3-month intervention consisted of 6 group sessions, 2 weeks apart, with an introductory session delivered by the district health program team of the government of Assam and Piramal Swasthya. This team were trained by the Piramal Swasthya’s experts, who received training on the Voeding Leeft’s reverse diabetes program principles using telemedicine.^30^ Patients in the intervention cluster met physically every fortnight. To boost effective communication and adherence to the intervention and to foster strong peer-to-peer relationships between members, an in-group peer educator was purposefully selected within each intervention cluster. The dietary intervention consisted of increasing the protein-carbohydrate ratio in meals, a 3-meal regimen with no snacking in between, and avoiding processed and high glycemic-load food. The participants were supported with a recipe booklet in the Assamese language that contained 30 simple recipes consisting of low-cost, locally available food items. These recipes factored in dietary principles of the intervention protocol and served as an adjunct tool to daily decision making. Nutritional tracking was performed using a diary in which participants logged their meal timings and contents. This was tracked by a patient-peer and guided by an expert nutritionist from Assam using group video-consultations. This was transferred to the study management team, who also advised on healthy dietary interventions. Participants were encouraged to choose an exercise befitting their daily chores. Physical activity advice included daily walking/aerobic workout for 15 - 30 minutes before and after the two main meals. Participants were encouraged to keep track of daily exercise in a provided diary. Additional advice included relaxation and stress reduction, maintaining a regular sleep routine and tobacco and alcohol cessation. The participants were informed about signs of hypoglycemia and supported with a safety plan. Participants received a glucose meter and were trained and prompted to measure their glucose levels on a 4-point curve (fasting and before each meal) for the first month to receive instant feedback. Henceforth, these 4-point curves were done on a need basis as decided by the specialist doctor who was also part of the program.

#### Participant Data

We obtained detailed sociodemographic data from each participant. Additionally, detailed lifestyle-related data, including dietary intake using a validated food frequency questionnaire (FFQ) were obtained.^32^ Quality of life was assessed using the World Health Organisation (WHO) Quality of Life (QOL) and General Health Questionnaire (GHQ-12) in various domains of health.^35^ The answers on the Likert scale were converted to numerical values. Biochemical parameters were measured in 8-hour fasting serum for glucose, HbA1c, and lipid parameters (total, low-density lipoprotein (LDL) cholesterol, high-density lipoprotein (HDL) cholesterol and triglycerides). The baseline data for all clusters were collected at the start of the study in September 2022, and outcome data were obtained at the end of the study in January 2023. Data regarding blood glucose and dietary parameters were also obtained fortnightly at cluster-level meetings.

### Outcomes

Primary outcomes were (a) diabetes remission defined as HbA1c <6.5% without diabetes medications as recommended by the UK NHS,^23^ and (b) differences in end-of-the-study HbA1c and fasting glucose levels in intervention and standard-care groups. Secondary outcomes were differences in weight, body mass index, blood pressure (BP), quality of life scores, and lipid profile components. This is an open-label study, and participants, peer-educators, and health workers were aware of the randomization groups. The outcome analysts and data managers were unaware of the randomization status.

### Patient and public involvement

The patient peers and health workers at primary healthcare centres in Assam,^34^ were involved in the study design, conduct, and measurement of outcomes of the study. All the patients were involved in the conduct of the study and measurement of the outcomes. The participants were not involved in the preparation of the manuscript.

### Statistical analysis

To estimate the required number of participants, power calculations have been carried out to obtain a statistical significance of a change in one of the primary outcomes, HbA1c, using G-power.^36^ A minimum of 274 participants in total was derived from these calculations. However, due to the nature of this study, additional calculations were required to determine the sample size needed to achieve the desired power in a clustered format. The required sample size was calculated utilizing a correlation coefficient of 0.01 for the co-primary outcome (HbA1c),and included intra-cluster correlation correction. The equation used was:

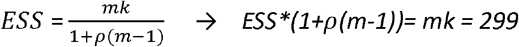

where, ESS = effective sample size (n=274), m = number of participants in a cluster (10 participants per cluster), mk = total number of participants and ρ = the correlation coefficient 0.02. To achieve the desired power, a minimum of 299 participants were included. Categorical data were compared with the Chi-Square test, and for the numerical data we used the non-parametric Mann-Whitney U-test. Differences between pre-test and post-test outcomes were assessed for median difference and 95% confidence intervals using the Custodial-Hodges-Lehmann nonparametric test. Analyses were conducted using IBM SPSS Statistics for Windows, version 27.0 (Armonk, NY, IBM Corp).

## RESULTS

The study flow diagram for the participants with type 2 diabetes is in Figure 1. 26 villages (clusters) were identified and were randomised to 13 clusters each for standard care and intervention. One cluster in standard care opted out due to an insufficient number of participants. Therefore, 160 type 2 diabetes patients in 12 clusters were in the standard care arm, and 193 patients in 13 clusters were in the intervention arm. (Figure 1). The number of participants who discontinued participation or were lost to follow-up is also in Figure 1.

**Figure 1.**
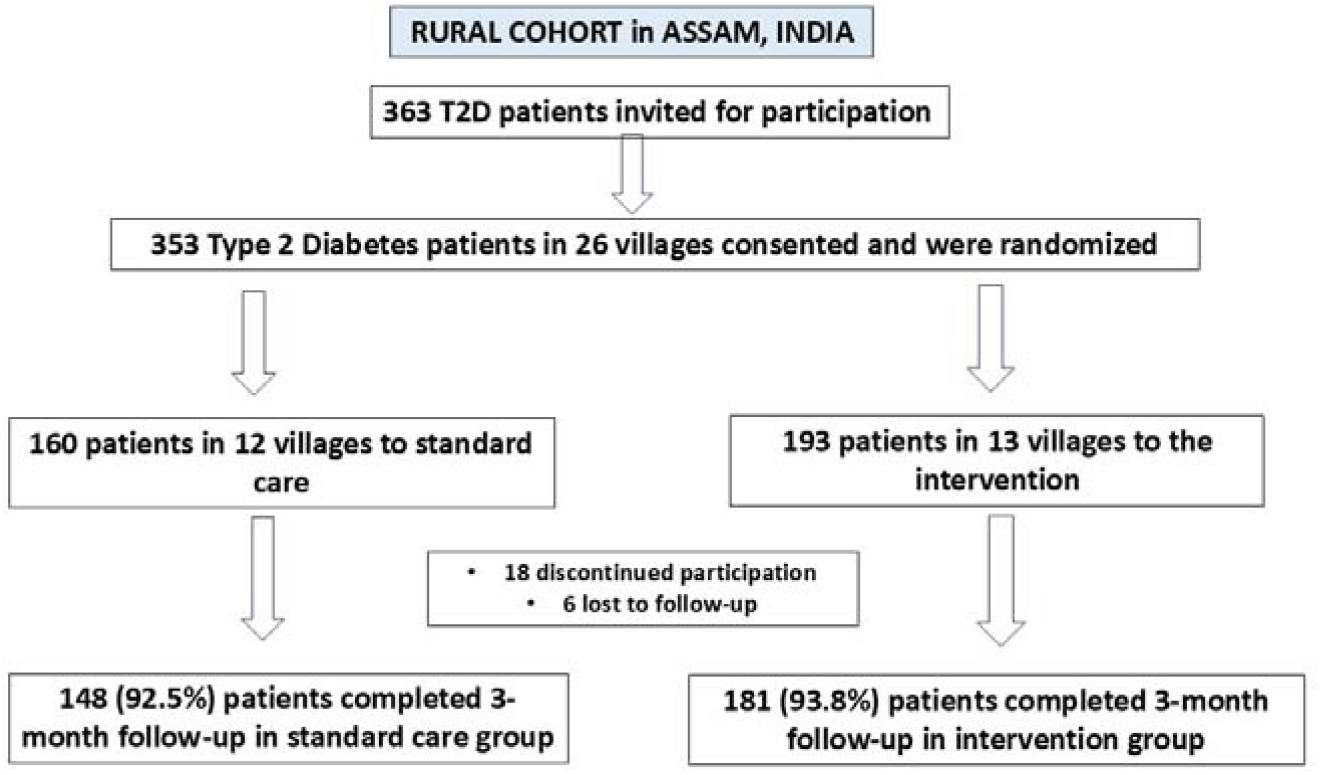
**Patient flow diagram in the study**

### Baseline data

The baseline characteristics of the study participants are in Table 1. Among participants in the standard-care and intervention groups the baseline sociodemographic, major lifestyle habits, clinical characteristics, and biochemical parameters were mostly identical. In the intervention group, the mean body mass index was more and more patients were on diabetes medicines. The health assessments in various domains-physical, social and environmental-were performed using multiple questionnaires for quality of life, physical health and independence, psychological health, social relationships, environmental health and overall health (WHO-general health questionnaire-12). No differences are observed at baseline in the standard-care and intervention groups (Supplementary Table 1). Qualitative FFQ data did not show a significant difference in consumption of healthy foods-whole grain cereals, fruits, vegetables, nuts, polyunsaturated and monounsaturated fats and dairy in standard-care vs intervention groups (Supplementary Table 2).

**Table 1.** Baseline characteristics of participants in standard care and intervention groups.

|  | Standard care (n=160) | Intervention (n=193) | P value |
| --- | --- | --- | --- |
| **Sociodemographic variables** |  |  |  |
| Age (median, IQR) | 50.0(43.0-50.7) | 50.0(42.0-54.5) | 0.125 |
| Age <40 yr | 26(16.3) | 32(16.6) | 0.996 |
| Age 40-60 | 107(66.9) | 142(73.6) | 0.566 |
| Age>60 | 27(16.9) | 19(9.8) | 0.087 |
| Men | 77(48.1) | 96(49.7) | 0.762 |
| Women | 83(51.9) | 97(50.3) | 0.863 |
| Married | 144(90.0) | 185(95.9) | 0.083 |
| Uneducated or <12y | 143(89.4) | 176(91.2) | 0.911 |
| Graduates | 17(10.6) | 17(8.8) | 0.565 |
| Regularly employed | 37(23.1) | 28(14.5) | 0.038 |
| Sedentary employment | 26(16.2) | 18(9.3) | 0.084 |
| Family size $\geq 5$/family | 99(61.9) | 116(60.1) | 0.734 |
| Smoking | 23(14.4) | 22(11.4) | 0.404 |
| Smokeless tobacco | 58(36.3) | 85(44.0) | 0.138 |
| Alcohol abuse | 10(6.3) | 11(5.7) | 0.828 |
| Family history T2D | 49(30.6) | 89(46.1) | 0.003 |
| T2D duration, years, median(IQR) | 3.5(2.0-5.0) | 4.0(2.0-6.0) | 0.870 |
| Diabetes medicines |  |  |  |
| None | 0(0.0) | 25(13.0) | <0.001 |
| Oral antidiabetics (OAD) | 157(98.1) | 164(85.0) | 0.035 |
| OAD + herbal | 3(1.9) | 4(2.1) | 0.994 |
| **Physiological and biochemical variables (median, IQR)** |  |  |  |
| Weight, Kg | 58.0(50.0-65.0) | 60.0(53.5-67.5) | 0.069 |
| Body mass index, kg/M<sup>2</sup> | 22.8(20.7-25.4) | 23.8(22.0-26.4) | 0.007 |
| Waist circumference, cm | 90.0(84.2-95.7) | 91.0(85.0-97.0) | 0.361 |
| Fasting glucose, mg/dl | 206.0(141.5-302.0) | 211.0(160.5-314.0) | 0.284 |
| HbA1c, % | 8.4(6.9-10.4) | 9.2(7.0-11.2) | 0.061 |
| Systolic BP, mmHg | 130.0(120.0-140.0) | 130.0(119.5-140.0) | 0.149 |
| Diastolic BP, mmHg | 80.0(70.7-86.5) | 80.0(70.0-90.0) | 0.859 |
| Cholesterol, mg/dl | 149.0(125.2-168.7) | 158.0(124.5-176.5) | 0.320 |
| HDL cholesterol, mg/dl | 43.0(35.0-48.9) | 39.0(32.0-47.0) | 0.106 |
| LDL cholesterol, mg/dl | 89.8(62.2-115.7) | 80.0(61.0-107.0) | 0.132 |
| Triglyceride, mg/dl | 187.0(139.0-228.5) | 169.0(133.0-220.5) | 0.209 |
| **Qualitative health assessment (Likert scale)** |  |  |  |
| Quality of life | 3.97(1.85) | 4.64(2.09) | 0.894 |
| Physical health | 17.58 (3.75) | 18.18(3.59) | 0.661 |
| Psychological health | 13.38(4.09) | 14.51(4.72) | 0.786 |
| Social relationships | 6.03(2.55) | 6.84(2.94) | 0.992 |
| Environmental health | 16.05(6.31) | 17.76(7.61) | 0.811 |
| Overall health (GHQ-12) | 14.65(5.39) | 13.07(5.71) | 0.444 |
| Inter-group comparisons have been performed using the Chi-square test for categorical variables and Mann-Whitney U-test for continuous variables. BP blood pressure, GHQ general health questionnaire, HDL high density lipoprotein, IQR interquartile range, LDL low density lipoprotein, T2D type 2 diabetes |  |  |  |

### Follow-up outcomes

Diabetes remission has been defined as HbA1c <6.5% without any anti-diabetes medications.^23,37^ Among the rural participants following 3 months of the study, it was observed in 4 patients (2.7%) in the standard-care group vs 9 (5.1%) in the intervention group (p=0.249) (Figure 2). Co-primary and secondary outcomes are in Table 2. Comparisons of primary and secondary outcomes at 3 months show that in the intervention vs standard care groups, there were significantly lower median (25-75^th^ interquartile range, IQR) HbA1c 7.9% (IQR 7.0-8.7) vs 8.6% (IQR 7.6-9.8) (p<0.001) and fasting glucose 188.0 mg/dl (IQR 146.2-253.5) vs 210.0 mg/dl (IQR 166.0-282.0) (p=0.001) (Figure 2).

**Table 2.** Comparisons of primary and secondary outcomes in standard care and intervention groups at 3-month follow-up.

|  | Standard care (n=148) | Intervention (n=181) | P value |
| --- | --- | --- | --- |
| **Primary outcome** |  |  |  |
| Diabetes remission | 4 (2.7%) | 9 (5.0%) | 0.249 |
| **Co-primary outcomes** |  |  |  |
| HbA1c (%) | 8.6(7.6-9.8) | 7.9(7.0-8.7) | <0.001 |
| Glucose fasting (mg/dl) | 210.0(166.0-282.0) | 188.0(146.2-253.5) | 0.001 |
| **Secondary outcomes** |  |  |  |
| Weight, Kg | 59.0(51.0-65.0) | 60.0(54.0-68.0) | 0.160 |
| Body mass index, kg/M<sup>2</sup> | 23.2(21.2-25.3) | 23.8(22.1-26.4) | 0.006 |
| Waist circumference, cm | 90.0(86.0-96.0) | 90.0(85.0-96.0) | 0.685 |
| Systolic BP, mmHg | 135.0(120.0-144.0) | 129.5(120.0-140.0) | 0.003 |
| Diastolic BP, mmHg | 80.0(78.0-90.0) | 80.0(74.0-81.0) | 0.100 |
| **Cholesterol, mg/dl** | 160(138.0-180.0) | 152.0(131.0-184.7) | 0.441 |
| **HDL cholesterol, mg/dl** | 42.0(34.0-47.0) | 42.5(36.0-51.0) | 0.075 |
| **LDL cholesterol, mg/dl** | 77.6(55.0-103.8) | 78.0(59.0-96.7) | 0.932 |
| **Triglycerides (mg/dl)** | 176.0(110.0-277.0) | 175.5(130.0-232.2) | 0.732 |
| Inter-group comparisons have been performed using the Chi-square test for categorical variables and Mann-Whitney U-test for continuous variables. BP blood pressure, HDL high density lipoprotein, IQR interquartile range, LDL low density lipoprotein, T2D type 2 diabetes |  |  |  |

**Figure 2.**
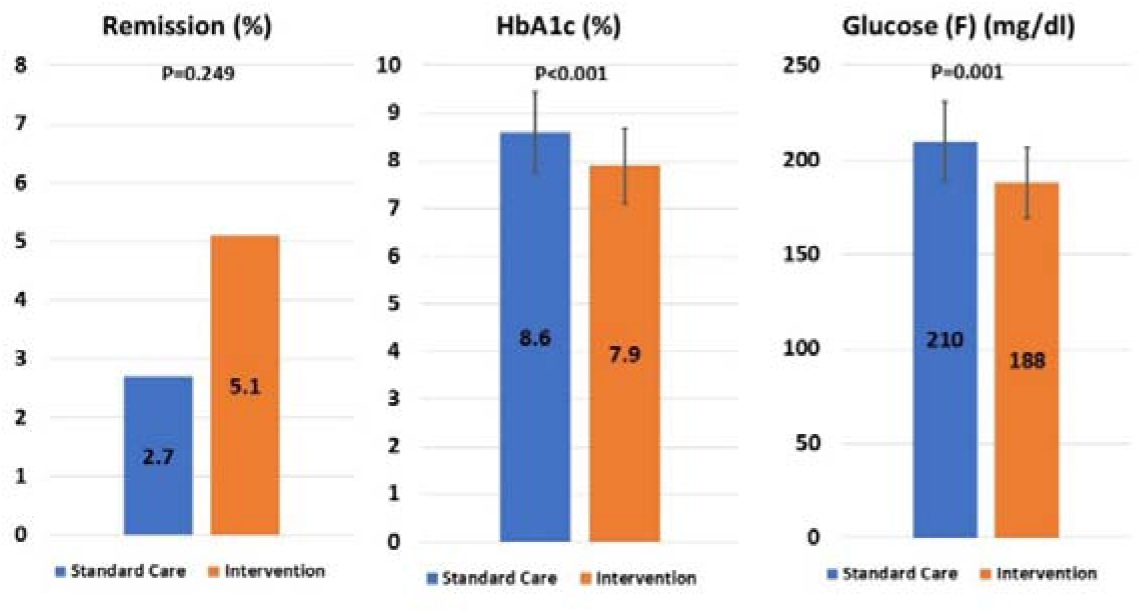
**Comparison of primary outcomes at the end of the study in standard care and intervention groups. Remission rates are in per cent, HbA1c and fasting (F) glucose are medians and 25^th^-75^th^ interquartile range.**

Comparison of secondary outcomes showed that in intervention vs standard care groups the systolic BP (129.5, 120.0-140.0 mmHg vs 135.0, 120-144.0 mmHg, p=0.003) was significantly lower. No difference was observed in other parameters (Table 2).

Median change in physiological and biochemical parameters from baseline to the end of the study in standard-care and intervention groups is shown in Table 3. Within-group before-after analyses show that in the standard care group median HbA1c increased from 8.3 (7.2-9.6%) to 8.6 (7.6-9.8%) (p=0.003) while in the intervention group it decreased from 9.5 (8.1-11.1%) to 7.9 (7.0-8.7%) (p<0.001); fasting glucose in standard care group increased from 201.0 (152.0-266.0 mg/dl) to 210 (166.0-282.0 mg/dl) (p=0.014) and declined significantly in intervention group from 232.0 (181.7-319.7 mg/dl) to 188.0 (146.2-253.5 mg/dl) (p<0.001). Significant declines in secondary outcomes of weight, body-mass index, waist circumference, and systolic and diastolic BP were also observed in the intervention group (Table 3).

**Table 3.**
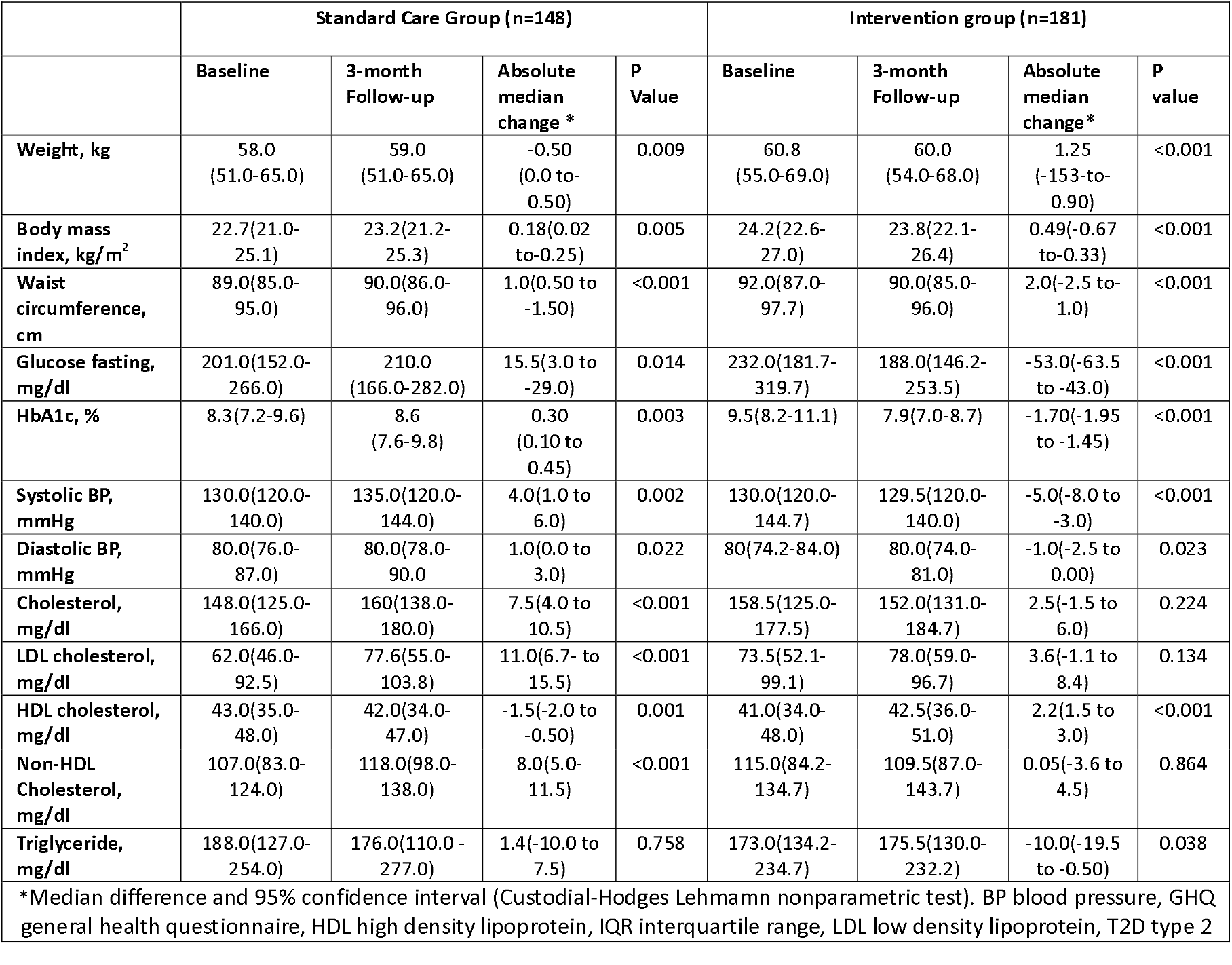
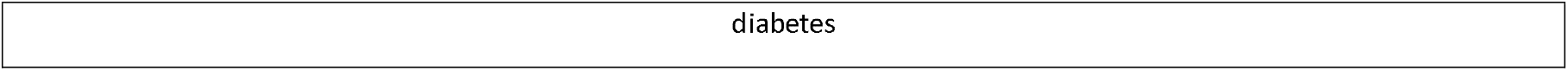
Change in physiological and biochemical variables at 3-month follow-up in standard care and intervention groups.

Change in various lifestyle factors at the end of the study period is shown in Figure 3. There is a significant decline in smokeless tobacco consumption in both control and intervention groups, with more in the intervention group.

**Figure 3.**
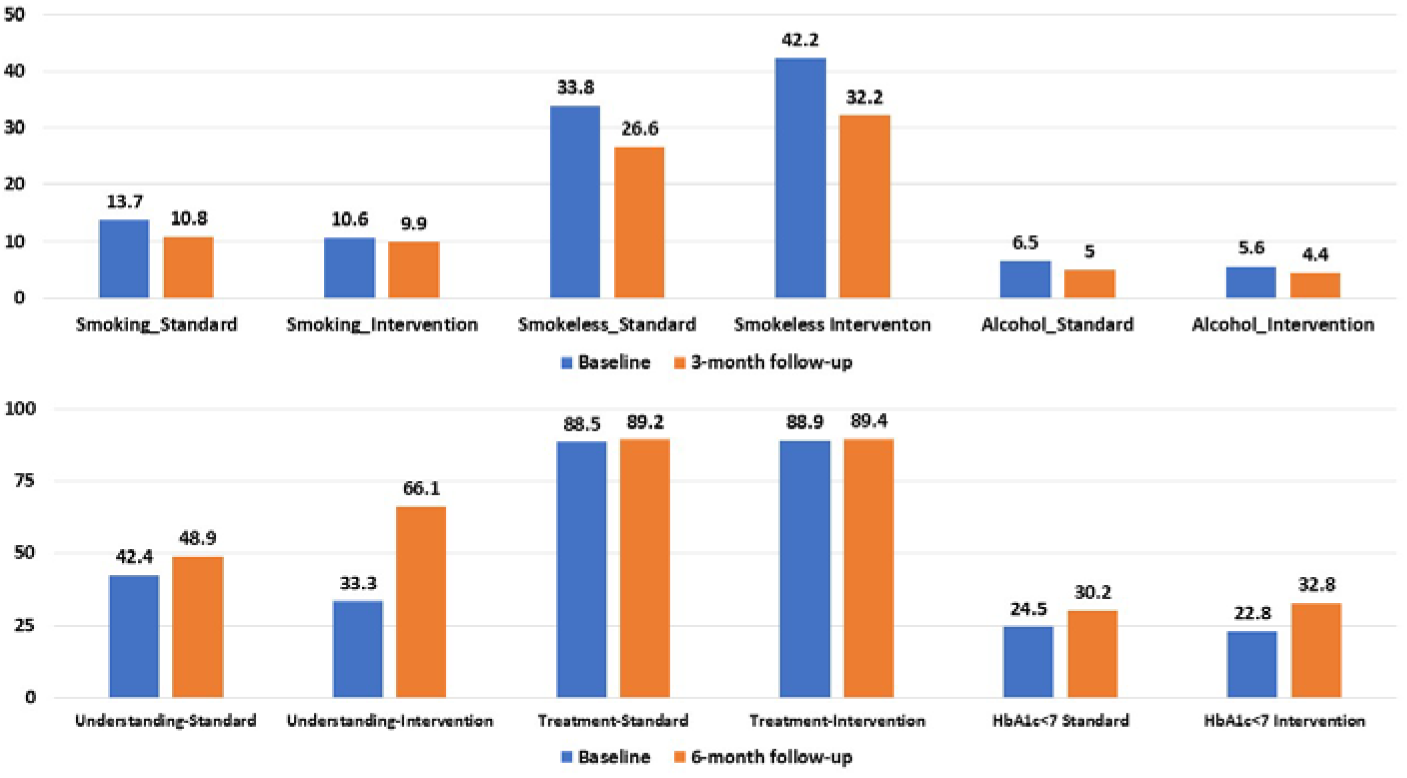
**Per cent differences in lifestyle factors, and understanding of diabetes and its treatment in standard-care and intervention groups at baseline and 3-month follow-up**

Changes in various domains of quality of life at 3-month follow-up showed significant improvements in the intervention arm for multiple parameters of well-being (p<0.05) (Figure 4).

**Figure 4.**
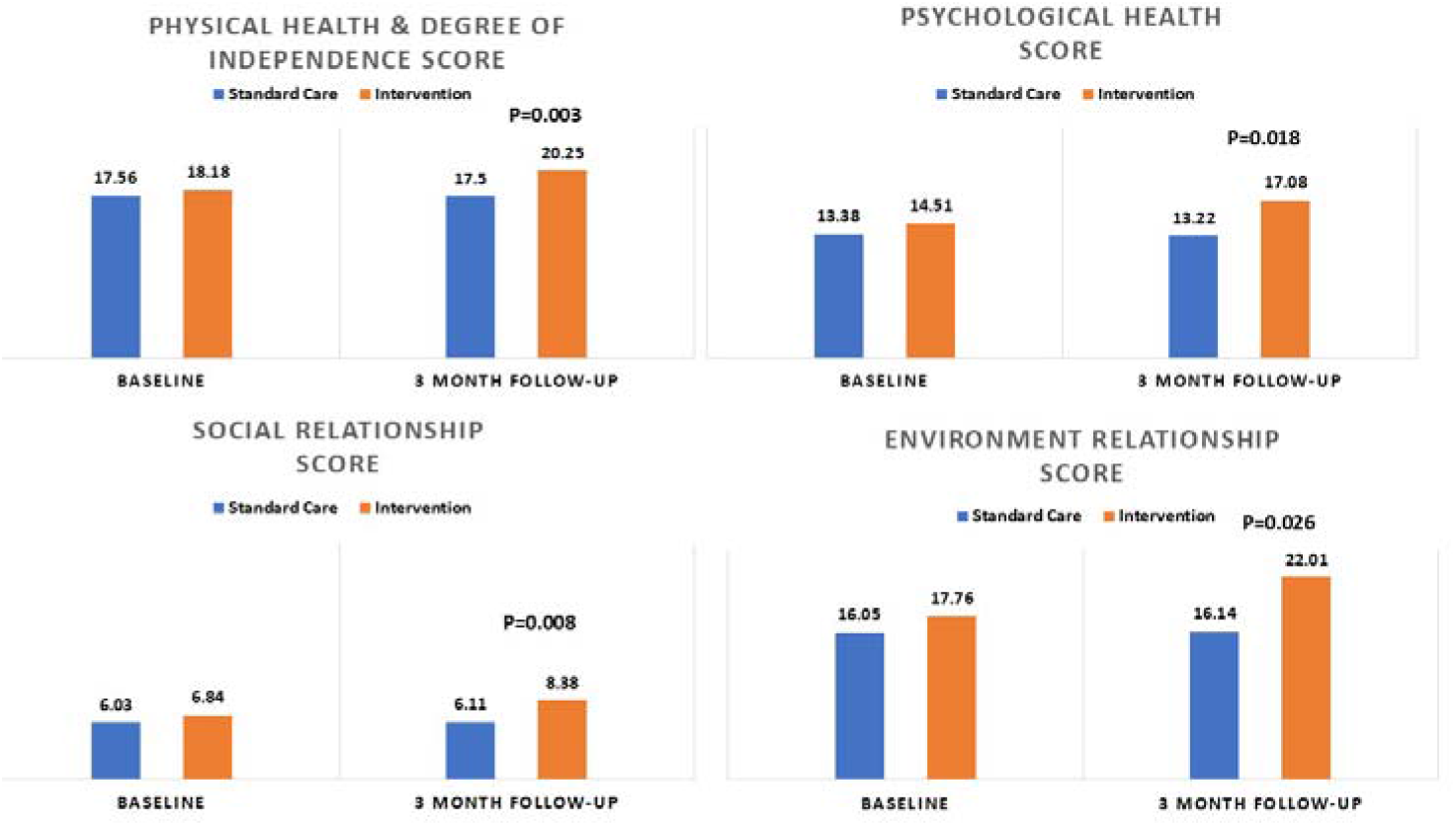
**Participants in the intervention arm reported significantly better outcomes for various qualitative scores, including for physical health and degree of independence, psychological health, social relationships and environmental relationships compared to the standard care.**

Various domains of the WHO quality of life questionnaire significantly improved in participants receiving the intervention as compared to the standard care, namely, physical health and degree of independence (−0.08 vs 2.08, scale 7-35, p=0.003); psychological health (−0.08 vs 2.56, scale 6-30, p=0.018); social relationship (0.08 vs 1.58, scale 3-15, p=0.026); and environmental health (0.08 vs 4.37, scale 8-40, p=0.008). Qualitative data on various parameters on the importance of diabetes, its treatment and control were also obtained and show significant improvement in the understanding of diabetes in the intervention group (Figure 4).

## DISCUSSION

This cluster-randomised study in rural locations in India shows that a patient-peer delivered, healthcare worker and technology-supported intervention for type 2 diabetes remission and control led to significant changes in diabetes control among study participants. Diabetes remission was achieved in a low proportion. Larger and longer-term studies using these interventions are required to confirm the effectiveness of the present strategy for the remission and control of type 2 diabetes in India and other lower-income and low-income countries.

Multi-component approaches for diabetes control include healthy behaviour change with a focus on individual-level patients. A review of 7205 abstracts and 13 detailed studies by Darcy et al reported that the focus on social determinants of health (poverty alleviation, education, better health systems) is important for behaviour change in diabetes.^37^ It was recommended that the multifactorial components should include the coordination of multi-disciplinary health care teams, in-person self-care classes, group activities, incorporation of peer-leaders, the development of community partnerships, economic relief and built-environment support to achieve better diabetes control. In the present intervention, we used most of these components along with telemedicine-based technological support.^32^ In the present study, the social determinants-based approach with a focus on underserved rural populations worked well, and suggests that our intervention is more suited for such participants. This is important because more than 80% of diabetes patients in India do not have access to continuous care as suggested by guidelines.^4,38^

The present study utilised a multicomponent intervention of patient-peer led, community-based model combining patient volunteers, healthcare workers, nutritionists, and clinicians, and telemedicine to deliver lifestyle advice for glycemia control in type 2 diabetes patients in rural India. The significant reduction in HbA1c, fasting glucose, and body weight observed in the study underscores the potential of peer-led, multicomponent lifestyle interventions in addressing the growing diabetes burden in underserved areas. This finding aligns with India’s National Programme for Prevention & Control of Non-Communicable Diseases,^7^ which aims to strengthen chronic care through community-based, task-sharing models. Incorporating such community-driven models into primary care, especially through Health and Wellness Centres in India,^7^ could offer a scalable, affordable strategy to improve diabetes outcomes in low-access regions. While this was a short-term study, the use of trained patient volunteers makes the approach both feasible and adaptable within existing public health frameworks and helps close the diabetes care gap in rural India,^4,5^ where more than two-thirds of the population resides and faces limited healthcare access.^7^

Strategies to induce diabetes remission mainly focus on weight loss using a variety of techniques-calorie restriction, low-carbohydrate diets, intermittent fasting and a plethora of patented diets.^41^ The DIRECT trial in the UK showed diabetes remission in more than a third of the participants,^21,22^ using a dietary intervention with focus on micronutrient complete low-energy diet typically in the form of soups and shakes to achieve diabetes remission, followed by food reintroduction and weight maintenance using a structured supprt.^21^ This approach was successful in 34% of patients for up to 5 years.^42^ DIRECT-like studies in other parts of the world and other clinical trials have failed to achieve this level of success.^25^ A systematic review and meta-regression analysis of 22 randomised controlled trials reported that diabetes remission was dependent on weight loss: for every 1% decrease in bodyweight, the probability of reaching complete remission increased by 2·17% (95% CI 1·94-2·40) and reaching partial remission increased by 2·74% (2·48-3·00).^43^ A significant heterogeneity was observed among clinical trials. A diabetes remission study using dietary weight management intervention in South Asians in the UK has reported rates identical to Caucasians.^44^ Metabolic surgery in patients with type 2 diabetes is associated with significant weight loss and a higher incidence of diabetes remission as compared to medical interventions.^45^ In the present study, the achieved median weight loss was modest (−1.25kg or −2.1%), and unsurprisingly, there was negligible diabetes remission. The study period was short, and hence we cannot compare our study with the longer-duration studies.

The study has several limitations that include short duration of study, different sample sizes in intervention and control arms of rural populations, differential availability and access to care in rural settings, level of adherence to dietary intervention, minimal weight loss and external validity. We used a stringent criteria to define type 2 diabetes remission, which has been suggested by recent consensus statements.^41^ Using a more liberal definition (HbA1c <7.0 with reduced drug intake) that was used in our previous study,^32^ would have resulted in remission rates of more than 10%. The follow-up duration is also shorter (3 months) than in previous studies (12-24 months); hence, the results are not comparable. Our study, on the other hand, is important as it shows that a multicomponent strategy for diabetes remission is feasible in India, especially in rural populations where diabetes is already epidemic.^3^ An important finding is a very high enrolment rate and low drop-out in the study populations (Figure 1). This is much higher than in most previous studies and shows the importance of our program in underserved populations of India. Quality of care and the presence of multiple influencers with disparate advice are important problems in Indian healthcare.^4^ Resolution is possible only when coordinated healthcare services are provided.

In conclusion, in this cluster-randomized study, we demonstrate the feasibility and efficacy of a diet and lifestyle intervention program for better control of type 2 diabetes and its possible remission, tailored to the local needs in rural populations in India. This multicomponent intervention showed significantly improved HbA1c, fasting glucose, and quality of life outcomes in rural populations. Larger and long-term studies are required to confirm the findings of the present study and to implement this intervention across diverse populations as a similar multicomponent tailored approach, facilitated with artificial intelligence algorithms,^46^ could greatly help in confronting the type 2 diabetes epidemic in India and other lower-middle and low-income countries.

## DECLARATIONS

### Contributors

JvdB, A-M E-S, EC, SKBH, ACB, LB, SL, LH, RG, and Voeding Leeft initiated the study. SKBH, SG, RV, VL, SL and KKS contributed to project administration, supervision and analyses. SKBH, SG, A-M E-S, EC, RV, ACB, LB, HP, GH, EV, HY, NvdZ, VL, SL and KKS supervised the study conduct. KKS, SKBH, SG, RV, NH, DG, MC, AV, CB, RB, SB, HB, VL, and SL contributed to the study with patient management. KKS, HY, and RG performed the statistical analyses. KKS, HY and RG wrote the first draft and revised all the subsequent drafts. All the authors reviewed and provided critical inputs to the manuscript. SKBH and KKS are responsible for the overall content and are the guarantors.

### Patient and public involvement

The patient peers were involved in the study design, conduct, and measurement of outcomes of the study. All the patients were involved in the conduct of the study and outcome measures. The participants or the public were not involved in the preparation of the manuscript. Data dissemination and scale-up strategy shall be implemented following publication of the study outcomes.

### Patient consent for publication

Individual informed consent was obtained from all the participants. Patient consent for publication: Not applicable.

### Ethics approval

The study involved human participants and was approved by the Institutional Ethics Committee, Piramal Swasthya Management and Research Institute, Hyderabad, India. All the patients provided informed consent prior to participation in the study.

### Provenance and peer review

Not commissioned; externally peer reviewed.

### Data availability statement

Data are available with the principal investigators at reasonable request. The Joep Lange Institute did have access to the collected data. Data are not publicly available.

## SUPPLEMENTARY DATA

**Supplementary Table 1.** Quality of life questionnaires at baseline and 3-month follow-up in intervention and standard-care groups.

|  | Lifestyle variables: (Likert scale, 0 to 10 scores) | Intervention group (N=180) |  |  | Standard care group (N=139) |  |  |
| --- | --- | --- | --- | --- | --- | --- | --- |
|  |  | Baseline | Follow-up | P value | Baseline | Follow-up | P value |
| 1 | How many times do you eat each day (Meals) | 3.0(2.0-3.0) | 3.0(2.0-3.0) | 0.014 | 2.0(2.0-3.0) | 2.0(2.0-3.0) | 0.297 |
| 2. | How many times do you eat each day (Snacks/ Tea/Coffee ) | 2.0(1.0-3.0) | 2.0(1.0-2.0) | <0.001 | 2.0(1.0-2.0) | 2.0(1.0-3.0) | 0.003 |
| 3 | Total score in (Almost every day/100% of the time) | 2.0(0.0-6.0) | 3.5(1.0-6.0) | 0.371 | 3.0(0.0-5.0) | 3.0(1.0-5.0) | 0.793 |
| 4. | Total score in (Often (Abt. 50%-75% of the time)) | 4.0(2.0-6.0) | 4.0(1.0-6.0) | 0.003 | 4.0(2.0-5.0) | 4.0(2.0-5.0) | 0.928 |
| 5. | Total score in (At times (Abt 25%-50% of the time) | 3.0(1.0-4.0) | 3.0(1.0-4.0) | 0.148 | 2.0(1.0-4.0) | 2.0(1.0-4.0) | 0.952 |
| 6. | Total score in (Almost never (Abt. 0%-25 % of the time)) | 2.0(0.0-4.0) | 3.0(1.0-5.0) | 0.037 | 2.0(1.0-3.0) | 2.0(1.0-3.0) | 0.576 |
| 7. | How would you rate your quality of life? | 4.0(3.0-5.0) | 5.0(4.0-8.0) | <0.001 | 3.0(3.0-4.0) | 4.0(3.0-4.0) | 0.633 |
| 8. | How satisfied are you with your health? | 4.0(3.0-5.0) | 4.0(4.0-8.0) | <0.001 | 3.0(3.0-4.0) | 3.0(3.0-4.0) | 0.285 |
| 9. | To what extent do you feel that physical pain prevents you from doing what you need to do? | 3.0(2.0-5.0) | 4.0(3.0-5.0) | 0.039 | 3.0(2.0-4.0) | 3.0(3.0-4.0) | 0.987 |
| 10. | How much do you need any medical treatment to function in your daily life? | 3.0(2.0-4.0) | 3.0(3.0-5.0) | 0.014 | 3.0(3.0-4.0) | 3.0(3.0-4.0) | 0.997 |
| 11. | How much do you enjoy life? | 4.0(3.0-5.0) | 4.0(4.0-8.0) | <0.001 | 4.0(3.0-4.0) | 4.0(3.0-4.0) | 0.245 |
| 12. | To what extent do you feel your life to be meaningful? | 4.0(3.0-5.0) | 4.0(4.0-8.0) | <0.001 | 4.0(3.0-4.0) | 4.0(3.0-4.0) | 0.445 |
| 13 | How well are you able to concentrate? | 4.0(3.0-5.0) | 4.0(4.0-8.0) | <0.001 | 4.0(3.0-4.0) | 4.0(3.0-4.0) | 0.424 |
| 14. | How safe do you feel in your daily life? | 4.0(3.0-5.0) | 4.0(4.0-8.0) | <0.001 | 3.0(3.0-4.0) | 3.0(3.0-4.0) | 0.061 |
| 15. | How healthy is your physical environment? | 4.(3.0-5.0) | 4.5(4.0-8.0) | <0.001 | 3.0(3.0-4.0) | 4.0(3.0-4.0) | 0.923 |
| 16. | Do you have eOUGH energy for everyday life? | 4.0(3.0-5.0) | 4.0(3.0-8.0) | <0.001 | 4.0(3.0-4.0) | 3.0(3.0-4.0) | 0.638 |
| 17 | Are you able to accept your bodily appearance? | 4.0(3.0-5.0) | 4.5(4.0-8.0) | <0.001 | 3.0(3.0-4.0) | 3.0(3.0-4.0) | 0.546 |
| 18 | Have you eOUGH money to meet your needs? | 4.0(3.0-5.0) | 4.0(4.0-7.0) | <0.001 | 3.0(3.0-5.0) | 3.0(3.0-4.0) | 0.078 |
| 19 | How available to you is the information that you need in your day-to-day life? | 4.0(3.0-5.0) | 4.0(3.0-8.0) | <0.001 | 3.0(2.0-4.0) | 4.0(3.0-4.0) | 0.944 |
| 20 | To what extent do you have the opportunity for leisure activities? | 3.0(3.0-5.0) | 4.0(3.0-7.0) | <0.001 | 4.0(3.0-4.0) | 3.0(3.0-4.0) | 0.056 |
| 21 | How well are you able to get around? | 4.0(3.0-5.0) | 4.0(4.0-8.0) | <0.001 | 3.0(2.0-4.0) | 4.0(3.0-5.0) | 0.159 |
| 22 | How satisfied are you with **your sleep?** | 4.0(3.0-5.0) | 4.0(4.0-8.0) | <0.001 | 4.0(3.0-4.0) | 4.0(3.0-5.0) | 0.862 |
| 23 | How satisfied are you with your ability to perform your daily living activities? | 4.0(3.0-5.0) | 4.0(4.0-8.0) | <0.001 | 4.0(3.0-5.0) | 4.0(3.0-4.0) | 0.923 |
| 24 | How satisfied are you with your capacity for work? | 4.0(3.0-5.0) | 4.0(4.0-8.0) | <0.001 | 4.0(3.0-4.0) | 4.0(3.0-4.0) | 0.118 |
| 25 | How satisfied are you with yourself ? | 4.0(3.0-5.0) | 5.0(4.0-8.0) | <0.001 | 4.0(3.0-5.0) | 4.0(3.0-4.0) | 0.373 |
| 26 | How satisfied are you with your persol relationships? | 4.0(3.0-5.0) | 5.0(4.0-8.0) | <0.001 | 4.0(3.0-5.0) | 4.0(3.0-5.0) | 0.687 |
| 27 | How satisfied are you with your sex life? | 4.0(3.0-5.0) | 4.0(4.0-7.0) | <0.001 | 4.0(3.0-5.0) | 4.0(3.0-5.0) | 0.187 |
| 28 | How satisfied are you with the support you get from your friends? | 4.0(3.0-5.0) | 5.0(4.0-8.0) | <0.001 | 3.0(2.0-4.0) | 4.0(3.0-5.0) | 0.588 |
| 29 | How satisfied are you with the conditions of your living place? | 4.0(3.0-5.0) | 5.0(4.0-8.0) | <0.001 | 4.0(3.0-5.0) | 4.0(3.0-5.0) | 0.550 |
| 30 | How satisfied are you with your access to health services? | 4.0(3.0-5.0) | 5.0(4.0-8.0) | <0.001 | 4.0(3.0-5.0) | 4.0(3.0-5.0) | 0.996 |
| 31 | How satisfied are you with your transport? | 4.0(3.0-5.0) | 5.0(4.0-8.0) | <0.001 | 4.0(3.0-5.0) | 4.0(3.0-5.0) | 0.634 |
| 32 | How often do you have negative feelings such as blue mood, despair, anxiety, depression? | 3.0(2.0-5.0) | 3.0(2.0-5.0) | 0.605 | 3.0(3.0-4.0) | 3.0(3.0-5.0) | 0.020 |

**Supplementary Table 2.** Dietary intake according to food-frequency questionnaire at baseline and 3-month follow-up.

|  | Lifestyle variables: (Likert scale, scores 0 to 10) | Intervention group (N=180) |  |  | Standard care group (N=139) |  |  |
| --- | --- | --- | --- | --- | --- | --- | --- |
|  |  | Baseline | Follow-up | P value | Baseline | Follow-up | P value |
| 1 | How many times do you eat each day: number of meals | 3.0(2.0-3.0) | 3.0(2.0-3.0) | 0.014 | 2.0(2.0-3.0) | 2.0(2.0-3.0) | 0.297 |
| 2. | How many times do you eat each day: Snacks/ Tea/Coffee | 2.0(1.0-3.0) | 2.0(1.0-2.0) | <0.001 | 2.0(1.0-2.0) | 2.0(1.0-3.0) | 0.003 |
| 3 | Total score for almost every day (100% of time) | 2.0(0.0-6.0) | 3.5(1.0-6.0) | 0.371 | 3.0(0.0-5.0) | 3.0(1.0-5.0) | 0.793 |
| 4. | Total score for often (50-75% of time) | 4.0(2.0-6.0) | 4.0(1.0-6.0) | 0.003 | 4.0(2.0-5.0) | 4.0(2.0-5.0) | 0.928 |
| 5. | Total score for sometimes (25-50% of time) | 3.0(1.0-4.0) | 3.0(1.0-4.0) | 0.148 | 2.0(1.0-4.0) | 2.0(1.0-4.0) | 0.952 |
| 6. | Total score for almost never (0-25 % of time) | 2.0(0.0-4.0) | 3.0(1.0-5.0) | 0.037 | 2.0(1.0-3.0) | 2.0(1.0-3.0) | 0.576 |

